# Quantitative G6PD point-of-care test can be used reliably on cord blood to identify male and female newborns at increased risk of neonatal hyperbilirubinaemia: a mixed method study

**DOI:** 10.1101/2022.07.03.22277173

**Authors:** Germana Bancone, Mary Ellen Gilder, Elsie Win, Gornpan Gornsawun, Penporn Penpitchaporn, Paw Khu Moo, Laypaw Archasuksan, Nan San Wai, Sylverine Win, Ko Ko Aung, Ahmar Hashmi, Borimas Hanboonkunupakarn, Francois Nosten, Verena I Carrara, Rose McGready

## Abstract

**Introduction:** New point-of-care (POC) quantitative G6PD testing devices developed to provide safe radical cure for P. vivax malaria may be used to diagnose G6PD deficiency in newborns at risk of severe neonatal hyperbilirubinaemia, improving clinical care, and preventing related morbidity and mortality. Methods: We conducted a mixed-methods study analyzing technical performance and usability of the “STANDARD G6PD” Biosensor when used by trained midwives on cord blood samples at two rural clinics on the Thailand-Myanmar border.

**Results:** In 307 cord blood samples, the Biosensor had a sensitivity of 1.000 (95%CI 0.859-1.000) and a specificity of 0.993 (95% CI 0.971-0.999) as compared to gold standard spectrophotometry to diagnose G6PD deficient newborns using a receiving operator characteristic (ROC) analysis-derived threshold of ≤4.8IU/gHb. The Biosensor had a sensitivity of 0.727 (95%CI: 0.498-0.893) and specificity of 0.933 (95%CI: 0.876-0.969) for 30-70% activity range in females using ROC analysis-derived range of 4.9 to 9.9IU/gHb. These thresholds allowed identification of all G6PD deficient neonates and 80% of female neonates with intermediate phenotypes.

Need of phototherapy treatment for neonatal hyperbilirubinaemia was higher in neonates with deficient and intermediate phenotypes as diagnosed by either reference spectrophotometry or Biosensor.

Focus group discussions found high levels of learnability, willingness, satisfaction, and suitability for the Biosensor in this setting. The staff valued the capacity of the Biosensor to identify newborns with G6PD deficiency early (“We can know that early, we can counsel the parents about the chances of their children getting jaundice”) and at the POC, including in more rural settings (“Because we can know the right result of the G6PD deficiency in a short time. Especially for the clinic which does not have a lab”). Conclusions: The Biosensor is a suitable tool in this resource-constrained setting to identify newborns with abnormal G6PD phenotypes at increased risk of neonatal hyperbilirubinaemia.

## INTRODUCTION

Pathologically increased levels of bilirubin during the first week of life, i.e. neonatal hyperbilirubinaemia (NH), are common and dangerous for the developing brain. The most severe form of NH, kernicterus, causes neurological sequelae in >80% of neonates (56/100,000 live births globally, [1]). Every year, an estimated twenty-four million newborns are at risk of NH-related adverse outcomes with three-quarters of mortality occurring in sub-Saharan Africa and South Asia [1,2]. These preventable deaths and disabilities disproportionally affect neonates where universal health care and treatment options are scarce, if not absent [3].

Several genetic and clinical factors influence the timing and evolution of NH, including G6PD deficiency, ABO blood group incompatibility, prematurity/low birth weight and sepsis [4]. Early identification of these risk factors can dramatically improve neonatal clinical management during the first days of life [5].

The enzymatic defect of G6PD deficiency, caused by mutations on the X-linked G6PD gene, is a known risk factor for increased levels of bilirubin after birth and it is associated with susceptibility to drug-induced haemolysis [6]. Risk of severe NH is increased in both deficient and heterozygous newborns with abnormal phenotypes [7-9] and universal neonatal screening of G6PD deficiency is supported by WHO in populations where more than 3-5% of males are affected [10].

G6PD deficiency is particularly prevalent among neonates from tropical regions [11], where clinical care is often provided in a non-tertiary hospital or clinic context. Knowledge of G6PD status by medical staff and parents can aid in avoiding potentially haemolytic antibiotics or other agents (such as naphthalene), improved follow-up, and heightened awareness of signs and symptoms of severe NH.

G6PD deficiency is very common among the Karen and Burman population along the Thailand-Myanmar border (9-18% in males, [12]) where it is associated with an increased risk to develop NH requiring phototherapy both in G6PD deficient (over 4-fold [13]) and in heterozygous females (over 2-fold [5]) as compared to wild type genotype neonates. In a recent study, screening of G6PD by qualitative Fluorescent Spot Test (FST) on cord blood failed to identify almost 10% of G6PD deficient neonates [14].

Demonstrating usability of new quantitative Point-Of-Care (POC) G6PD diagnostic tests by locally trained clinical staff can inform clinical deployment in this setting and in other rural settings. This study assessed the technical performance and usability of the “G6PD STANDARD” (SD Biosensor, Korea) test when used by trained midwives in two clinics along the Thailand-Myanmar border.

## MATERIALS AND METHODS

### Study design

A mixed-methods study was conducted to evaluate both the technical performance of the “G6PD STANDARD” (SD Biosensor, Korea) test (henceforth “Biosensor”) and its usability by midwives in a non-tertiary setting. G6PD enzymatic activity and haemoglobin concentration measured by the device were compared to the gold standard reference spectrophotometric assay and haematology analyser, respectively. Performance of the G6PD fluorescent spot test (FST) currently used routinely at the point-of-care, was also compared to the reference and new test.

Following local staff training, user proficiency was assessed before study start; usability was explored using focus group discussions (FGD) at the end of the study.

### Study setting and population

The study was conducted in SMRU clinics situated along the Thailand-Myanmar border in Tak province (Thailand) where free antenatal care and birthing services are provided for migrant women of predominantly Karen and Burman ethnicity.

SMRU midwives come from the same population as the pregnant women and patients seeking care at SMRU clinics. The majority of midwives have primary or secondary education and receive clinical training on-site. Pregnant women attending SMRU clinics at Wang Pha (WPA) and Maw Ker Thai (MKT) were informed about the study at regular antenatal care visits in the 3^rd^ trimester. Informed consent procedures and eligibility assessments for mothers were completed before labour commenced. Eligibility of neonates was assessed immediately after delivery, and those born at an estimated gestational age (EGA) by ultrasound ≥35 weeks with no severe maternal complications at delivery and no severe neonatal illness were included. In order to allow laboratory analyses to be performed within 30 hours from collection, only neonates born during week days were included. For all neonates, indication for starting phototherapy treatment followed the recommendations of the UK NICE guidelines [15].

### Blood analyses for technical evaluation of Biosensor

Two milliliters of cord blood were collected into EDTA from the umbilical cord using an established SMRU SOP. An aliquot of anticoagulated blood was used by the midwives in the delivery room for the Biosensor following manufacturer’s instructions within one hour of collection (Appendix 1). Tests were repeated if the test result was an error or “HI” (a result obtained when G6PD activity is very high, outside the instrument analytic range). High-level and low-level Biosensor controls were run weekly or monthly (depending on availability) from April 2020 until May 2021.

An aliquot of anticoagulated blood was analysed by G6PD fluorescent spot test (FST) at the clinical laboratory. The remaining blood was stored at 4°C until shipment to the central SMRU laboratory on the same day.

Gold standard reference testing for G6PD and haemoglobin were performed by spectrophotometric assay and haematology analyser (with complete blood and reticulocyte counts), respectively, at the SMRU central laboratory.

G6PD spectrophotometric assay was performed using Pointe Scientific kits (assay kit # G7583-180, Lysis Buffer # G7583-LysSB). Kinetic determination of G6PD activity at 340 nm was performed using a SHIMAZU UV-1800 spectrophotometer with temperature controlled cuvette compartment (30°C). Samples were analysed in double and mean activity was expressed in IU/gHb using the Hb concentration obtained by complete blood count analysis. The final result was calculated using manufacturer’s Temperature Control Factor of 1.37. Two controls (Normal, Intermediate or Deficient; Analytic Control Systems, Inc. USA) were analysed at every run and results compared to expected ranges provided by manufacturer. Complete blood count was performed using a CeltacF MEK-8222K haematology analyser (Nihon Kohden, Japan). Three-levels quality controls were run every day and device maintenance and calibration were performed regularly. Reticulocytes were analysed by microscopy after staining with supervital staining Crystal Violet.

Buffy coat recovered from whole blood after centrifugation was stored at -20°C for later DNA extraction using standard columns kit (Favorgen Biotech, Taiwan). Genotyping for G6PD common mutations was performed through established SOPs [16]. Mahidol mutation was analysed in all samples. Other mutations were only analysed in phenotypically deficient or intermediate samples (G6PD < 9.31IU/gHb by reference test) with wild type or heterozygote Mahidol genotypes. Viangchan, Chinese-4, Kaiping, Canton, Union and Mediterranean were analysed first and full gene sequence was performed if none of these mutations were found.

### Biosensor training, user proficiency and usability assessment

Midwives of WPA and MKT SMRU clinics were trained for use of Biosensor and were eligible to participate in the usability component of the study following informed consent. Two to four training sessions were provided at each clinic in the local language by an experienced laboratory technician (author LA). The sessions lasted from 1 to 2 hours and included a short introduction about the test, a practical demonstration using imitation blood, and supervised use of the biosensor by each midwife. Midwives were allowed to practice the procedure the week following the training prior to taking a user proficiency test. The proficiency test was administered by author LA in the local language and it consisted of a questionnaire (modified from a questionnaire developed by PATH (https://www.finddx.org/wp-content/uploads/2020/09/PATH_STANDARD-G6PD-User-Competency-Assessment-quiz_08oct19.pdf) and direct observation of two consecutive tests. Midwives were asked to explain out-loud their actions while performing the first test. The proficiency test was analysed by authors GB and GG and midwives who scored <85% were re-trained before study start. A visual aid with all critical steps of the procedure was printed and available in the delivery room during the study. The usability component of the study followed the conceptual framework for acceptance and use of a rapid diagnostic test for malaria proposed by Asiimwe et al. [17] that evaluates 6 components: learnability, willingness, suitability, satisfaction, efficacy, and effectiveness. The focus group discussions (FGD) specifically focused on 4 main themes of learnability, willingness, satisfaction, and suitability. Due to COVID, only two of the planned six total FGD were conducted. The midwives were grouped by their seniority, with senior and junior midwives together, and midwife assistants in a separate group in order to encourage honest and open conversation. One researcher (KKA) facilitated the FGD while an experienced assistant took notes; both were fluent in Burmese and Karen languages used in the FGD. Immediately following the FGD, research staff debriefed and noted main themes of the discussion. FGDs were audio-recorded and subsequently translated and transcribed in English. Two researchers (MG and GB) independently analysed the transcript using thematic analysis based on the pre-set framework [17] using Taguette (a free and open access qualitative data analysis software, https://joss.theoj.org/papers/10.21105/joss.03522) and confirmed findings with the KKA. Face-to-face meeting and exchange of notes allowed for triangulation between the researchers.

### Blood analysis for assessment of neonatal hyperbilirubinaemia

Routine clinical care for newborns included at least one total serum bilirubin (TSB) test before discharge (around 48h of life) using capillary blood measured on-site by the rapid quantitative bilirubinometer BR-501 (Apel Co. Ldt, Japan).

### Sample size and statistical analyses

The expected prevalence of G6PD deficiency in the population living at the border is 9-18% in males and 2-4% in female [12, 16] corresponding to approximately 20-30% heterozygous females, 60% of whom have intermediate activity [18]. Assuming that the proportion of females and males in the neonate population is 50%, 9% were expected to be G6PD deficient and 7% to be G6PD intermediate. In order to obtain 95% CI of the limits of agreement within 0.5 SD of the difference, about 31 neonates with deficiency and 25 with intermediate phenotypes were needed, with a minimum total sample size of 350 samples.

Clinical data were double entered in MACRO and collated with laboratory data; data were analysed using SPSSv27.

Male median (MM) was calculated in all males with wild type genotypes in both the references spectrophotometric assay and the Biosensor. Deficiency was defined as enzymatic activity below 30% of MM by reference spectrophotometry and receiving operator characteristic (ROC)-derived 30% threshold by Biosensor; intermediate phenotypes were defined as enzymatic activity between 30% and 70% of the MM or ROC-derived threshold.

Mean and standard deviation (SD) were reported for continuous variables. Categorical variables were compared by Chi-squared test and ANOVA. Bland-Altman plot was used to inspect correspondence between G6PD activity detected by Biosensor compared to the spectrophotometry assay [19]. Correlation was assessed using Pearson’s coefficient of correlation and Interclass Correlation Coefficient (ICC). Area under the curve (AUC) of the ROC curve [20] was calculated at different activity thresholds to analyse clinical performances (i.e. sensitivity and specificity) of the Biosensor. Cohen’s Kappa coefficient was calculated for categories of phenotypes identified by Biosensor and spectrophotometry.

For analysis of haematologic features and risk of neonatal hyperbilirubinaemia, neonates gestational ages assessed by ultrasound were categorized as ≤38 and >38 weeks according to epidemiologic studies conducted previously in the same population [21].

Statistical significance was assessed at the 5% level.

### Patient and Public Involvement statement

At the outset of the study, the research team engaged the local population through a local ethics and research advisory committee, the Tak Province Community Advisory Board, Thailand. This group is comprised of community leaders, and were asked to advise on study design, process, and outcomes of interest, and subsequently approved the study (TCAB201904).

## RESULTS

A total of 331 cord blood samples were collected between April 2020 and November 2021; six were clotted and excluded from all analysis. Of the remaining 325 samples, 257 (79%) were collected in MKT clinic and 68 in WPA clinic, in 166 (51%) female and 159 male neonates. Mean (SD) of estimated gestational age of newborns was 39.1 (1.0) weeks.

### General haematologic characteristics

As expected for this specimen, haematological characteristics of cord blood (Table 1) show higher white blood cell count, haemoglobin concentrations, reticulocyte counts and larger cellular volumes compared to adult blood. Reticulocyte counts and red cell distribution width were higher in neonates <38 weeks gestational age (P=0.02 and P=0.01 respectively) while the other indexes did not differ by gestational age groups.

**Table 1.**
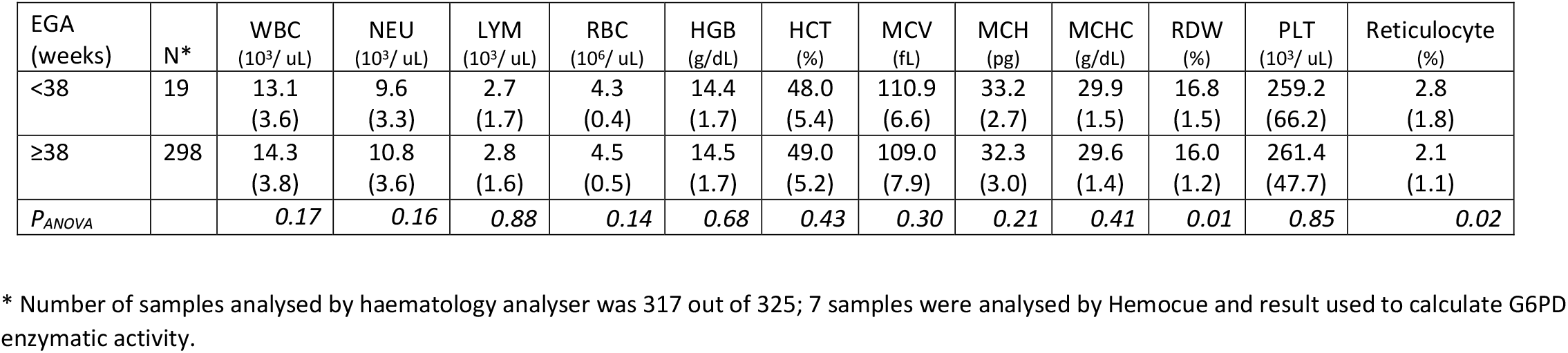
Haematologic characteristics of cord blood samples according to newborn gestational age. Results are shown as mean (SD)

| EGA<br>(weeks) | N* | WBC<br>(10<sup>3</sup>/ uL) | NEU<br>(10<sup>3</sup>/ uL) | LYM<br>(10<sup>3</sup>/ uL) | RBC<br>(10<sup>6</sup>/ uL) | HGB<br>(g/dL) | HCT<br>(%) | MCV<br>(fL) | MCH<br>(pg) | MCHC<br>(g/dL) | RDW<br>(%) | PLT<br>(10<sup>3</sup>/ uL) | Reticulocyte<br>(%) |
| --- | --- | --- | --- | --- | --- | --- | --- | --- | --- | --- | --- | --- | --- |
| <38 | 19 | 13.1<br>(3.6) | 9.6<br>(3.3) | 2.7<br>(1.7) | 4.3<br>(0.4) | 14.4<br>(1.7) | 48.0<br>(5.4) | 110.9<br>(6.6) | 33.2<br>(2.7) | 29.9<br>(1.5) | 16.8<br>(1.5) | 259.2<br>(66.2) | 2.8<br>(1.8) |
| ≥38 | 298 | 14.3<br>(3.8) | 10.8<br>(3.6) | 2.8<br>(1.6) | 4.5<br>(0.5) | 14.5<br>(1.7) | 49.0<br>(5.2) | 109.0<br>(7.9) | 32.3<br>(3.0) | 29.6<br>(1.4) | 16.0<br>(1.2) | 261.4<br>(47.7) | 2.1<br>(1.1) |
| *P*<sub>ANOVA</sub> |  | 0.17 | 0.16 | 0.88 | 0.14 | 0.68 | 0.43 | 0.30 | 0.21 | 0.41 | 0.01 | 0.85 | 0.02 |

\* Number of samples analysed by haematology analyser was 317 out of 325; 7 samples were analysed by Hemocue and result used to calculate G6PD enzymatic activity.

### G6PD genotypes

A total of 26 hemizygous mutated males (21 Mahidol, 2 Kaiping, 1 Viangchan, 1 Coimbra, 1 Orissa), 3 homozygous mutated females (Mahidol), 34 heterozygous females (32 Mahidol, 1 Canton, 1 Viangchan) and 262 wild type (129 females and 133 males) were found. Overall allelic frequency of all mutated alleles was 13.4%. The distribution of G6PD activity by spectrophotometry and biosensor associated with different genotypes are shown in Figures 1 and Supplementary Tables 1 and 2.

**Figure 1.**
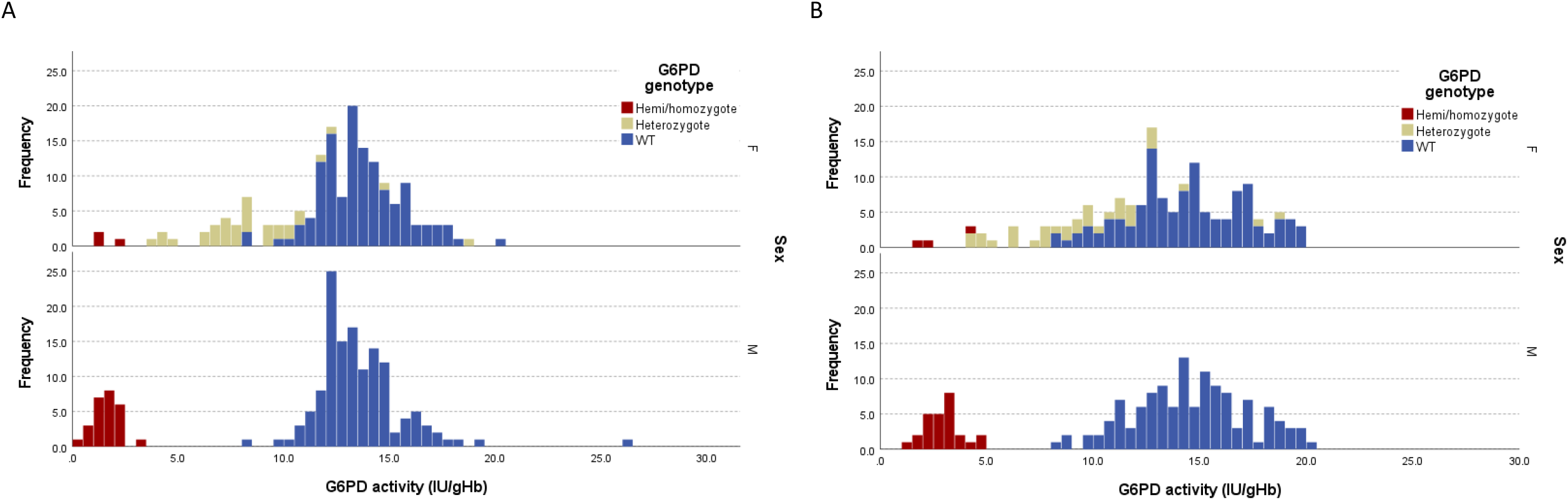
Distribution of G6PD enzymatic activity from cord blood samples detected by gold standard spectrophotometry assay (A) and Biosensor (B) according to sex and genotype

### Fluorescent spot test

The poor performances of the FST in cord blood were confirmed here, with the FST failing to identify 23% (7/30) of deficient neonates and 100% of the intermediate females (22/22; Table 2).

**Table 2.**
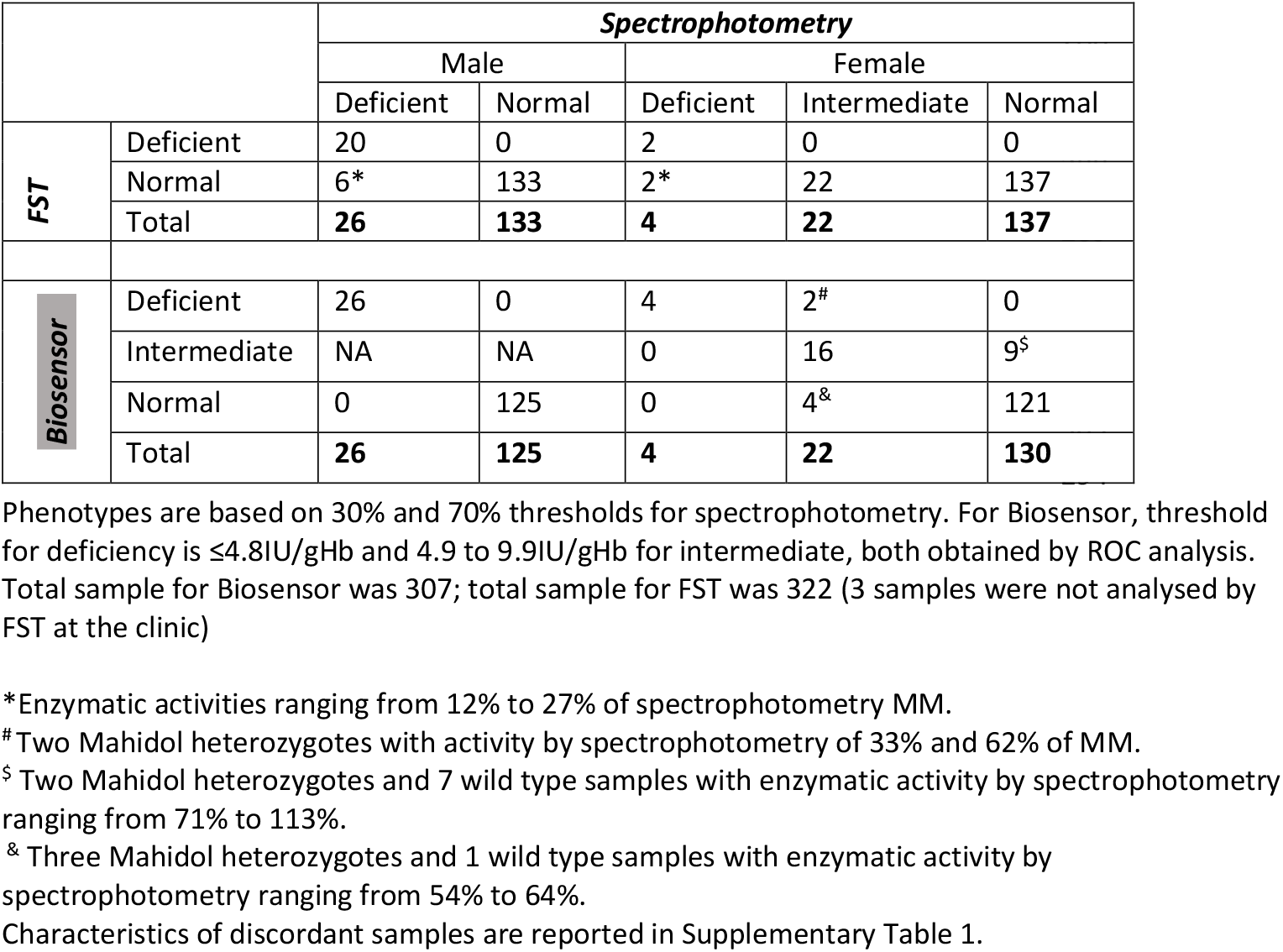
Diagnostic performance of FST and Biosensor as compared to gold standard spectrophotometry.

### Technical evaluation of Biosensor

#### Male medians by reference spectrophotometric assay and Biosensor

MM G6PD activity by spectrophotometer was 13.3 IU/gHb giving a 30% threshold of 4.0 IU/gHb for diagnosis of deficiency; intermediate activity (30-70%) in females ranged between 4.1 and 9.3 IU/gHb. The cord blood-specific 30% spectrophotometric threshold identified all the hemizygous male and homozygous female newborns (Figure 1A).

MM of G6PD activity by Biosensor calculated on 307 samples was 14.4 IU/gHb giving a 30% threshold of 4.3 IU/gHb for diagnosis of deficiency. Intermediate activity (30-70%) in females ranged between 4.4 and 10.1 IU/gHb (Figure 1B).

In 7% of cases (23/325), the Biosensor provided an initial result of “HI” activity without a numeric value. Of the 19 samples retested, 14 had “HI” results again and 5 samples had an activity ranging from 17.3 to 20.0 IU/gHb; all samples with initial or confirmed “HI” results were normal by spectrophotometry and had a wild type genotype. Overall, 18 samples (5.5% of the total) did not have a final numeric result by Biosensor but would have been considered “normal”, according to the spectrophotometric assay.

#### Biosensor performance

Biosensor performance was assessed for 307/325 samples that yielded numeric results. The mean (±1.96SD) difference in enzymatic activity between Biosensor and spectrophotometry was 1.05 IU/gHb (LoA: -3.52 to 5.62 IU/gHb) as represented in the Bland-Altman plot in Figure 2A. A very strong correlation between enzymatic activity by Biosensor and reference spectrophotometry was observed (Pearson’s r=0.855, p<0.001; ICC=0.905, p<0.001).

**Figure 2.**
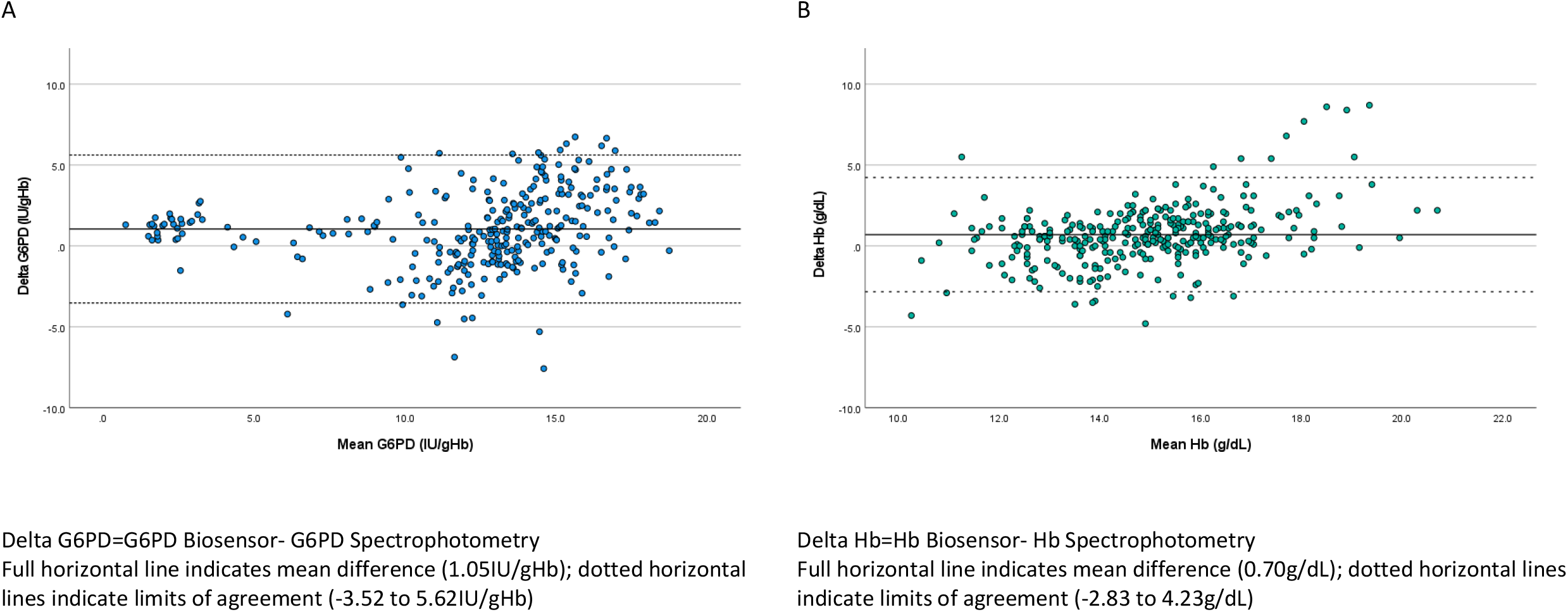
Bland Altman plot of G6PD activity (A) and haemoglobin levels (B) in cord blood comparing gold standard spectrophotometry to Biosensor

The mean (±1.96SD) difference in Hb between the Biosensor and haematology analyser was 0.70 g/dL (LoA: -2.83 to 4.23 g/dL) (Figure 2B). A moderate correlation between Hb levels by Biosensor and haematology analyser was observed (Pearson’s r=0.637, p<0.001; ICC=0.728, p<0.001).

Area under the curve (AUC) of the ROC analysis (Figure 3A) of the 30% threshold was 0.999 (95%CI: 0.997-1.000); ROC analysis showed that 30% of Biosensor MM (4.3IU/gHb) was associated with sensitivity of 0.931 (95%CI: 0.758-0.988) and specificity of 0.989 (95%CI: 0.966-0.997) while a threshold of 4.8IU/gHb had a sensitivity of 1.000 (95%CI: 0.859-1.000) and a specificity of 0.993 (95% CI: 0.971-0.999). This second threshold was therefore used for the subsequent analyses.

**Figure 3.**
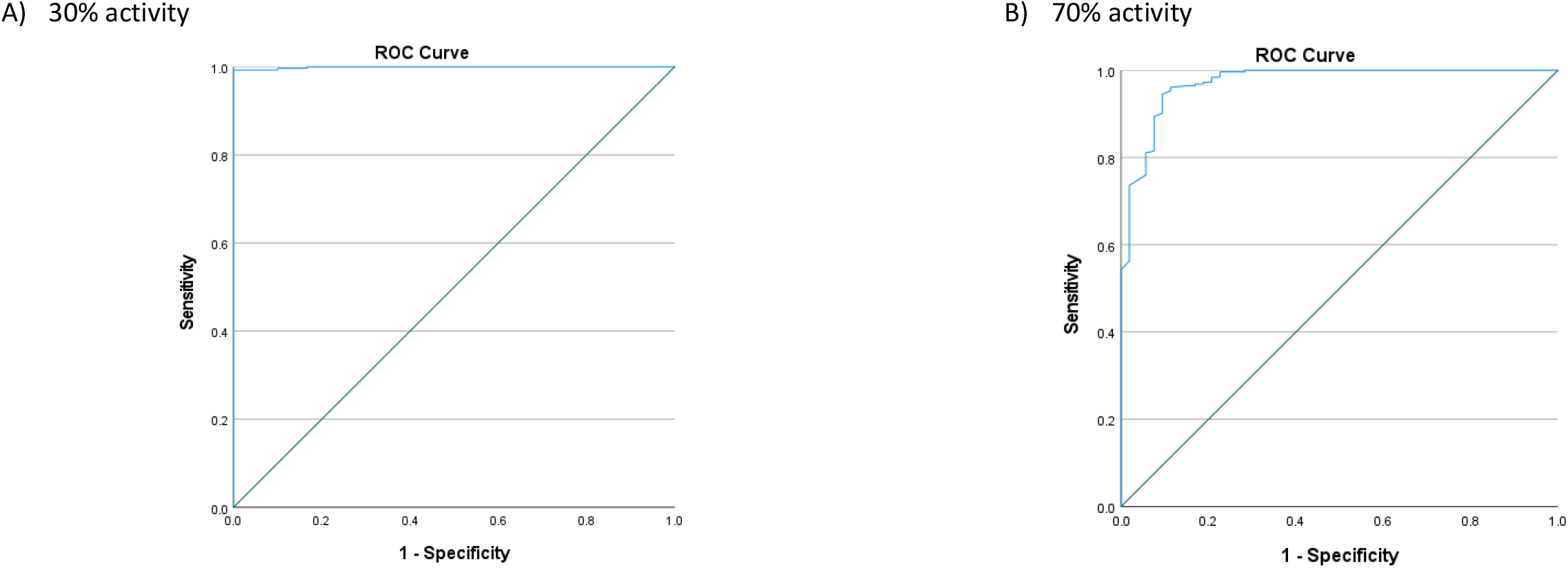
Receiver Operating Characteristic curve of Biosensor for 30% activity (A) and 70% activity (B) thresholds.

AUC of the ROC analysis (Figure 3B) for the 70% threshold was 0.972 (95%CI: 0.949-0.994) and ROC analysis showed that a threshold of 9.9IU/gHb had a better sensitivity and specificity as compared to the 70% of Biosensor MM (10.1 IU/gHb). The ROC-derived threshold had a sensitivity of 0.842 (95%CI: 0.716-0.921) and specificity of 0.984 (95%CI: 0.957-0.995) to identify samples with ≤70% activity and was used for subsequent analyses.

AUC of the ROC analysis for the range 30-70% activity was 0.935 (95%CI: 0.887-0.983); sensitivity and specificity for intermediate phenotypes in females were 0.727 (95%CI 0.498-0.893) and 0.933 (95%CI: 0.876-0.969) respectively based on ROC-derived thresholds as compared to 0.592 (95%CI: 0.390-0.770) and 0.953 (95%CI: 0.897-0.980) using Biosensor MM thresholds.

When comparing phenotypes defined according to the 30% and 70% thresholds of spectrophotometry and ROC-derived threshold for Biosensor (Table 2), the Biosensor correctly identified all deficient and normal males and all deficient females. In females, the Biosensor incorrectly identified 9% (2/22) of intermediate females (activity by spectrophotometry 33% and 62%) as deficient, and 7% (9/130) of phenotypically normal female neonates as intermediate (activity by spectrophotometer ranging from 71% to 113%). It also misdiagnosed 18% (4/22) of intermediate samples as normal. Of these 4 samples, 3 were Mahidol heterozygotes and 1 was a wild type and their enzymatic activity by spectrophotometry ranged from 54% to 64%. Cohen’s kappa coefficient was 0.841, p<0.001. Overall, the majority of samples with discordant results (11/15) were identified by the Biosensor as having a “worse” phenotype. Characteristics of the 15 samples with discordant results are reported in Supplementary Table 3.

No difference in results were observed by clinic (ICC=0.899, p<0.001 in MKT and ICC=0.930, p<0.001 in WPA) or user. In MKT clinic where the test was used over 20 months, a trend of larger absolute mean differences in activity (Biosensor - Spectrophotometry) were observed in the last 4-8 months of use as compared to the first 12 months (Supplementary Figure 1).

### Risk of neonatal hyperbilirubinaemia

Risk of neonatal hyperbilirubinemia by phenotype (determined by spectrophotometry) was assessed in term neonates (EGA≥38weeks). A significantly larger proportion of G6PD deficient neonates (29%) underwent phototherapy for treatment of NH as compared to G6PD normal (6%, RR[95%CI] =4.9 [2.3-10.5]; P<0.001). A larger proportion of female neonates with intermediate phenotypes (90% of whom were heterozygotes) required phototherapy (15%), although in this small cohort the difference did not reach statistical significance (RR[95%CI] =2.6 [0.8-8.1]; P=0.13; supplementary Table 4. Relative risk by quantitative phenotypes were similar to those already established by genotypes in the same population [5].

### Biosensor training, user proficiency and usability assessment

A total of 22 midwives in two clinics were initially trained and completed the users’ proficiency test, including 7 senior, 10 junior and 5 assistant midwives. Median (min-max) observed score from the questionnaire (max 7 points) and observed tests (max 18 points) was 22.1 (18-24.5). The median score did not differ by seniority: assistant 21.4 (18.0-23.5), junior 22.0 (19.3-24.5), senior 22.8 (21.0-24.5); most midwives (72%) had a score >21 points (>85% of maximum score). The most common mistakes in the questionnaire were on how to mix the blood and the buffer (pipetting 10 times *vs* shaking the buffer tube) and on volume of blood mixture to transfer into the device. On observation, the most common mistakes were failure to check the date on Biosensor screen and failure to check test expiry date (rated as minor mistakes as expired test strips are automatically recognized by the Biosensor and rejected).

Two focus group discussions were held in December 2021 in MKT clinic, four weeks after completion of the sample collection at that site; one FGD included 6 senior and junior midwives, and one included 6 assistant midwives. Discussions on satisfaction, learnability, willingness, and suitability and future use are summarized in Table 3. Overall satisfaction was high, although staff were concerned with invalid results, and found it challenging to dedicate one member of the team to perform the biosensor test in the delivery room in the busy postpartum period. In terms of learnability, the midwife assistants reported learning the device more easily, though some were anxious about missing steps. The senior staff were anxious about mistakes and clotted blood, and reported the need to refer to the instructions as a problem. Contrary to the positive expressions to keep using the device at the clinic, the midwives’ willingness to use the device was not high and they requested a dedicated staff to perform the test or the test to be done in the laboratory. In terms of suitability and future use, the midwives found the results clinically useful and a valuable diagnostic tool in both their setting and field clinics. However, they were concerned about neglecting clinical care while doing a laboratory test, the cost of the device, and emphasized the need for good training.

**Table 3.** Selected quotes by theme from focus group discussions.

| Theme | Quotes |
| --- | --- |
| A. Satisfaction | "It is very good for the children. It is good to know if the child has G6PD deficiency or not from birth. The advantage of the device is that it can detect the children without having to do a heel stick on the baby. On the other hand, there is an increase in work.... But now that we are good at using it, it's fine." [FGD1]<br>"Sometimes if someone is doing the test by using the device it means there are fewer staffs to be with mothers and babies which is not good." [FGD1] |
| B. Learnability | "After the one-time training, we had 1 or 2 times experiences practically. Then we can do it." [FGD2]<br>"I am really scared I will forget the steps." [FGD2]<br>"We have to look at the book very often, if not we forget the process of what to put and how to put it." [FGD1] |
| C. Willingness | "Facilitator: Yes. What do you think about keeping on using this device in the future?<br>Participant: Of course. It is good.<br>Participant: Yes, it is good. But if we can have a specific staff to do it then it will be better." [FGD2]<br>"To make changes, take out the blood and send it to the lab. Then only lab staff have to do that." [FGD1] |
| D. Suitability & Future Use | "Because we can know that early, we can have counseling with the parents about the chances of their children getting yellow skin. We can take time to counsel." [FGD1]<br>"Because we can know the right result of the G6PD deficiency in a short time. Especially for the clinic which doesn't have a lab then it is difficult to know the G6PD status. But with this device, they will only need to take a little blood from the baby and they can know the result of G6PD." [FGD2] |

## DISCUSSION

This is the first study to assess clinical performance and usability by locally trained health workers of the “STANDARD G6PD” Biosensor test for identification of G6PD deficient and intermediate phenotypes in cord blood. Current data, together with previously collected evidence from clinical trials in the same population [5], clearly indicate that newborn heterozygous girls with G6PD intermediate phenotypes, who are not identified by the FST, are at increased risk of NH and require phototherapy [7, 8]. The availability of a validated POC quantitative test such as the Biosensor and its inclusion in diagnostics guidelines for neonatal care at birth will allow identification of this group of neonates and better clinical care in several settings [22-25]. Together with other easy-to-use non-invasive tools for diagnosis of NH (e.g. Transcutaneous bilirubinometers), this study provides evidence that Biosensor could be used in non-tertiary rural settings for identification of neonates who need referral to higher levels of care. In settings where phototherapy is available, this study indicates that the Biosensor is a better option than FST to support clinical management of neonates. Technical performance of the Biosensor using ROC-derived threshold was comparable to that observed in adult blood in laboratory and field studies [26-29].

The phenotypic classification provided by the Biosensor was superior to the currently available qualitative test (FST) both for deficient and for intermediate phenotypes. Among intermediate phenotypes, 80% were identified as either deficient or intermediate, allowing a better identification of neonates at potential jaundice risk as compared to the currently used FST-based diagnosis [14, 30]. Poor performance of FST can be explained by the higher G6PD enzymatic activity at birth as compared to adulthood [31, 32]; this is probably the result of several haematological factors including younger red cell age, increased number of reticulocytes with higher G6PD activity [33, 34] and higher WBC count [28] as observed here. Importantly, because of higher enzymatic activity in cord blood, thresholds established in adult blood cannot be used to identify deficient or intermediate phenotypes by either spectrophotometry or Biosensor at birth and would have missed identification of 10% (3/29) deficient neonates (2/26 deficient males and 1/4 deficient females) and 86% (19/22) intermediate females.

Biosensor haemoglobin values had a moderate correlation with those assessed by automatic haematology analyser. Although cord (and neonatal) blood samples have higher haemoglobin levels and increased viscosity, Biosensor’s performance in measuring G6PD activity was not worse at higher haemoglobin levels.

While the Biosensor provided a numeric result in 94.5% of cases, in few cases an “error” message or a “HI” result was obtained which, according to the protocol, required re-analysis of the sample. Samples that tested “HI” were confirmed to be normal, both phenotypically by spectrophotometry and by genotype (all wild type). In routine practice it will not be needed to repeat the test in samples showing “HI” result should the manufacturers include this information in the instructions for use.

The usability component highlighted important themes to be taken into consideration for future use of the Biosensor at birth. The midwives have been involved in previous research regarding neonatal jaundice and appreciated the importance of early G6PD diagnosis to identify newborns most at risk of neonatal hyperbilirubinaemia and to facilitate optimal clinical care and parental counselling. The non-invasive nature of cord blood analysis was considered an advantage. In this setting, the SMRU midwives recommended that the test be performed by dedicated staff or by the available laboratory to assure appropriate clinical care is provided to the newborns and mothers; nevertheless, they estimated that in more rural contexts it may be appropriate for trained birth attendants to perform the test. Of note, midwives considered their reliance on reading the visual aid while performing the test (which is standard practice in laboratories) a weakness and this aspect might need to be taken into account when training clinic field staff. Usability results obtained here might not be generalizable to every other context but there are data being collected in several rural and community-based settings that corroborate ease of use of this device to guide malaria treatment after appropriate training [26, 35, 36].

Although midwives felt uncertain about properly conducting the test at the beginning of the study, the laboratory data showed highly accurate results in the first 12 months of use and very good results in the latter 8 months, supporting suitability of the test among health care workers without prior experience in diagnostics. Follow up studies should explore the causes of this slight decrease in quality over time which could be attributed to environmental or users’ factors as well as device durability over >1 year of use in tropical conditions.

### Limitations

A practical limitation of Biosensor testing on cord blood is the extra step needed to collect the blood with a syringe from the cord. A sampling device that collects a fixed volume of blood directly from the cord would streamline the process.

It is very likely that performance and reference ranges observed here in cord blood could apply to neonatal capillary or venous blood collected within the first 24 hours of life but this was not evaluated during the study.

The study was conducted in a period critically influenced by the COVID-19 pandemic. Travel restrictions resulted in a delayed study start, reduced enrolment in one clinic (WPA), and a protracted enrolment duration of the study overall. Fewer than planned FGD were conducted—including planned discussions at key time points during the study—and they occurred in a single clinical site providing a possibly narrower point of view on the usability topics explored. Additional staff stressors and human resource limitations due to COVID-19 and the political unrest in 2021 were not assessed but may have influenced the results of both the technical and usability components of the study.

### CONCLUSIONS

The “STANDARD G6PD” Biosensor is a reliable POC tool to support the perinatal care of newborns at higher risk of neonatal hyperbilirubinemia by demonstrating very high sensitivity in identification of deficient newborns and high sensitivity in identification of female newborns with intermediate activity. Its use by trained personnel in rural clinics and birthing centers with a high prevalence of G6PD deficiency, together with assessment of bilirubin levels before discharge, has the potential to avert disability and death from hyperbilirubinaemia.

Extending use of the Biosensor for newborn testing in countries where it is already deployed for malaria case management in resource-constrained settings [37], would provide a higher return on this investment. Use of Biosensor in populations with prevalent G6PD deficiency outside malaria endemic regions might increase the benefit-cost ratio of universal screening [38] in all settings [39].

## Data Availability

De-identified participant data are available from the Mahidol Oxford Tropical Medicine Data Access Committee upon request from this link: https://www.tropmedres.ac/units/moru-bangkok/bioethics-engagement/datasharing.The study was supported by a grant to GB by the Wellcome Trust Institutional Translational Partnership Awards: Thailand Major Overseas Programme (WT-iTP-2019/004). SMRU is supported by the Wellcome Trust [grant 220111]. For the purpose of Open Access, the authors have applied a CC BY public copyright licence to any author accepted manuscript version arising from this submission. The funders had no role in study design, data collection and analysis, decision to publish, or preparation of the manuscript.

## ACKNOWLEDGMENTS

The authors wish to thank all the mothers for theier collaboration and understanding; the study would not have been possible without the hard work and dedication of all SMRU staff involved, especially during such a difficult time of political unrest and COVID-19 pandemic. Acknowledgments are also extended to SD Biosensor for donating the devices and the tests for the study.

## ETHICAL APPROVAL

The study was approved by Oxford Tropical Research Ethics Committee, UK (OxTREC 532-19), the Mahidol University Faculty of Tropical Medicine Ethics Committee, Thailand (TMEC 19-048, MUTM 2019-080-02) and the Tak Province Border Community Ethics Advisory Board (TCAB201904). Written informed consent was obtained from literate mothers and midwives; a thumbprint was obtained in the presence of a literate witness for illiterate mothers.

## DATA AVAILABILITY STATEMENT

De-identified participant data are available from the Mahidol Oxford Tropical Medicine Data Access Committee upon request from this link: https://www.tropmedres.ac/units/moru-bangkok/bioethics-engagement/datasharing.

## REFERENCES

1. Bhutani VK, Zipursky A, Blencowe H, Khanna R, Sgro M, Ebbesen F, et al. Neonatal hyperbilirubinemia and Rhesus disease of the newborn: incidence and impairment estimates for 2010 at regional and global levels. Pediatric research. 2013;74 Suppl 1:86–100.

2. Bhutani VK, Vilms RJ, Hamerman-Johnson L. Universal bilirubin screening for severe neonatal hyperbilirubinemia. J Perinatol. 2010;30 Suppl:S6–15.

3. Olusanya BO, Kaplan M, Hansen TWR. Neonatal hyperbilirubinaemia: a global perspective. Lancet Child Adolesc Health. 2018;2(8):610–20.

4. Lauer BJ, Spector ND. Hyperbilirubinemia in the newborn. Pediatr Rev. 2011;32(8):341–9.

5. Bancone G, Gornsawun G, Peerawaranun P, Penpitchaporn P, Paw MK, Poe DD, et al. Contribution of genetic factors to high rates of neonatal hyperbilirubinaemia on the Thailand-Myanmar border. Plos Global Public Health. 2022.

6. Cappellini MD, Fiorelli G. Glucose-6-phosphate dehydrogenase deficiency. Lancet. 2008;371(9606):64–74.

7. Kaplan M, Hammerman C, Vreman HJ, Stevenson DK, Beutler E. Acute hemolysis and severe neonatal hyperbilirubinemia in glucose-6-phosphate dehydrogenase-deficient heterozygotes. The Journal of pediatrics. 2001;139(1):137–40.

8. Meloni T, Forteleoni G, Dore A, Cutillo S. Neonatal hyperbilirubinaemia in heterozygous glucose-6-phosphate dehydrogenase deficient females. British journal of haematology. 1983;53(2):241–6.

9. Molou E, Schulpis KH, Thodi G, Georgiou V, Dotsikas Y, Papadopoulos K, et al. Glucose-6-Phosphate Dehydrogenase (G6PD) deficiency in Greek newborns: the Mediterranean C563T mutation screening. Scandinavian journal of clinical and laboratory investigation. 2014;74(3):259–63.

10. WHO Working Group. Glucose-6-phosphate dehydrogenase deficiency. Bulletin of the World Health Organization. 1989;67(6):601–11.

11. Howes RE, Piel FB, Patil AP, Nyangiri OA, Gething PW, Dewi M, et al. G6PD deficiency prevalence and estimates of affected populations in malaria endemic countries: a geostatistical model-based map. PLoS medicine. 2012;9(11):e1001339.

12. Bancone G, Chu CS, Somsakchaicharoen R, Chowwiwat N, Parker DM, Charunwatthana P, et al. Characterization of G6PD Genotypes and Phenotypes on the Northwestern Thailand-Myanmar Border. PloS one. 2014;9(12):e116063.

13. Turner C, Carrara V, Aye Mya Thein N, Chit Mo Mo Win N, Turner P, Bancone G, et al. Neonatal intensive care in a Karen refugee camp: a 4 year descriptive study. PloS one. 2013;8(8):e72721.

14. Thielemans L, Gornsawun G, Hanboonkunupakarn B, Paw MK, Porn P, Moo PK, et al. Diagnostic performances of the fluorescent spot test for G6PD deficiency in newborns along the Thailand-Myanmar border: A cohort study. Wellcome open research. 2018;3:1.

15. Health NCCfWsCs. NICE. Neonatal jaundice: clinical guideline 2010.

16. Bancone G, Chowwiwat N, Somsakchaicharoen R, Poodpanya L, Moo PK, Gornsawun G, et al. Single Low Dose Primaquine (0.25mg/kg) Does Not Cause Clinically Significant Haemolysis in G6PD Deficient Subjects. PloS one. 2016;11(3):e0151898.

17. Asiimwe C, Kyabayinze DJ, Kyalisiima Z, Nabakooza J, Bajabaite M, Counihan H, et al. Early experiences on the feasibility, acceptability, and use of malaria rapid diagnostic tests at peripheral health centres in Uganda-insights into some barriers and facilitators. Implement Sci. 2012;7:5.

18. Bancone G, Gilder ME, Chowwiwat N, Gornsawun G, Win E, Cho WW, et al. Prevalences of inherited red blood cell disorders in pregnant women of different ethnicities living along the Thailand-Myanmar border. Wellcome open research. 2017;2:72.

19. Bland JM, Altman DG. Statistical methods for assessing agreement between two methods of clinical measurement. Lancet. 1986;1(8476):307–10.

20. Kumar R, Indrayan A. Receiver operating characteristic (ROC) curve for medical researchers. Indian pediatrics. 2011;48(4):277–87.

21. Thielemans L, Peerawaranun P, Mukaka M, Paw MK, Wiladphaingern J, Landier J, et al. High levels of pathological jaundice in the first 24 hours and neonatal hyperbilirubinaemia in an epidemiological cohort study on the Thailand-Myanmar border. PloS one. 2021;16(10):e0258127.

22. Aynalem YA, Mulu GB, Akalu TY, Shiferaw WS. Prevalence of neonatal hyperbilirubinaemia and its association with glucose-6-phosphate dehydrogenase deficiency and blood-type incompatibility in sub-Saharan Africa: a systematic review and meta-analysis. BMJ Paediatr Open. 2020;4(1):e000750.

23. Bernardo J, Nock M. Pediatric Provider Insight Into Newborn Screening for Glucose-6-Phosphate Dehydrogenase Deficiency. Clin Pediatr (Phila). 2015;54(6):575–8.

24. DelFavero JJ, Jnah AJ, Newberry D. Glucose-6-Phosphate Dehydrogenase Deficiency and the Benefits of Early Screening. Neonatal Netw. 2020;39(5):270–82.

25. Wong RJ, Montiel C, Kunda M, Stevenson DK, Bhutani VK. A novel point-of-care device for measuring glucose-6-phosphate dehydrogenase enzyme deficiency. Semin Perinatol. 2021;45(1):151356.

26. Alam MS, Kibria MG, Jahan N, Thriemer K, Hossain MS, Douglas NM, et al. Field evaluation of quantitative point of care diagnostics to measure glucose-6-phosphate dehydrogenase activity. PloS one. 2018;13(11):e0206331.

27. Pal S, Bansil P, Bancone G, Hrutkay S, Kahn M, Gornsawun G, et al. Evaluation of a Novel Quantitative Test for Glucose-6-Phosphate Dehydrogenase Deficiency: Bringing Quantitative Testing for Glucose-6-Phosphate Dehydrogenase Deficiency Closer to the Patient. The American journal of tropical medicine and hygiene. 2019;100(1):213–21.

28. Pal S, Myburgh J, Bansil P, Hann A, Robertson L, Gerth-Guyette E, et al. Reference and point-of-care testing for G6PD deficiency: Blood disorder interference, contrived specimens, and fingerstick equivalence and precision. PloS one. 2021;16(9):e0257560.

29. Zobrist S, Brito M, Garbin E, Monteiro WM, Clementino Freitas S, Macedo M, et al. Evaluation of a point-of-care diagnostic to identify glucose-6-phosphate dehydrogenase deficiency in Brazil. PLoS neglected tropical diseases. 2021;15(8):e0009649.

30. Kosaryan M, Mahdavi MR, Jalali H, Roshan P. Why does the Iranian national program of screening newborns for G6PD enzyme deficiency miss a large number of affected infants? Pediatric hematology and oncology. 2014;31(1):95–100.

31. Doherty AN, Kring EA, Posey YF, Maisels MJ. Glucose-6-phosphate dehydrogenase activity levels in white newborn infants. The Journal of pediatrics. 2014;164(6):1416–20.

32. Kaplan M, Abramov A. Neonatal hyperbilirubinemia associated with glucose-6-phosphate dehydrogenase deficiency in Sephardic-Jewish neonates: incidence, severity, and the effect of phototherapy. Pediatrics. 1992;90(3):401–5.

33. Ellis D, Sewell CE, Skinner LG. Reticulocyte enzymes and protein synthesis. Nature. 1956;177(4500):190–1.

34. Srivastava A, Evans KJ, Sexton AE, Schofield L, Creek DJ. Metabolomics-Based Elucidation of Active Metabolic Pathways in Erythrocytes and HSC-Derived Reticulocytes. Journal of proteome research. 2017;16(4):1492–505.

35. Engel N, Ghergu C, Matin MA, Kibria MG, Thriemer K, Price RN, et al. Implementing radical cure diagnostics for malaria: user perspectives on G6PD testing in Bangladesh. Malaria journal. 2021;20(1):217.

36. Gerth-Guyette E, Adissu W, Brito M, Garbin E, Macedo M, Sharma A, et al. Usability of a point-of-care diagnostic to identify glucose-6-phosphate dehydrogenase deficiency: a multi-country assessment of test label comprehension and results interpretation. Malaria journal. 2021;20(1):307.

37. Chu CS, Bancone G, Kelley M, Advani N, Domingo GJ, Cutiongo-de la Paz EM, et al. Optimizing G6PD testing for Plasmodium vivax case management: why sex, counseling, and community engagement matter. Wellcome open research. 2020;5:21.

38. Vidavalur R, Bhutani VK. Economic evaluation of point of care universal newborn screening for glucose-6-Phosphate dehydrogenase deficiency in United States. J Matern Fetal Neonatal Med. 2021:1–9.

39. Watchko JF, Kaplan M, Stark AR, Stevenson DK, Bhutani VK. Should we screen newborns for glucose-6-phosphate dehydrogenase deficiency in the United States? J Perinatol. 2013;33(7):499–504.

