## Supplementary Material for "Quantitative G6PD point-of-care test can be used reliably on cord blood to identify male and female newborns at increased risk of neonatal hyperbilirubinaemia: a mixed method study"

**Supplementary Figures and Tables**

**S Table 1.** G6PD enzymatic activity (IU/gHb) of cord blood by spectrophotometry according to genotype

| G6PD genotype | N | Mean | Std. Deviation | Minimum | Maximum |
|---|---|---|---|---|---|
| Hemizygote | 26 | 1.64 | 0.65 | 0.09 | 3.32 |
| Homozygote | 3 | 1.66 | 0.43 | 1.38 | 2.16 |
| Heterozygote | 34 | 8.55 | 2.97 | 3.54 | 18.89 |
| WT | 262 | 13.62 | 2.02 | 8.01 | 26.32 |
| Total | 325 | 12.02 | 4.14 | 0.09 | 26.32 |

**S Table 2.** G6PD enzymatic activity (IU/gHb) by Biosensor according to genotype

| G6PD genotype | N | Mean | Std. Deviation | Minimum | Maximum |
|---|---|---|---|---|---|
| Hemizygote | 26 | 2.87 | 0.81 | 1.4 | 4.6 |
| Homozygote | 3 | 2.70 | 1.23 | 1.8 | 4.1 |
| Heterozygote | 34 | 9.50 | 3.47 | 4.0 | 18.6 |
| WT | 244 | 14.46 | 2.72 | 8.1 | 20.0 |
| Total | 307 | 12.82 | 4.47 | 1.4 | 20.0 |

**S Figure 1.** Absolute difference in G6PD activity detected by Biosensor as compared to spectrophotometry over time (only MKT clinic)

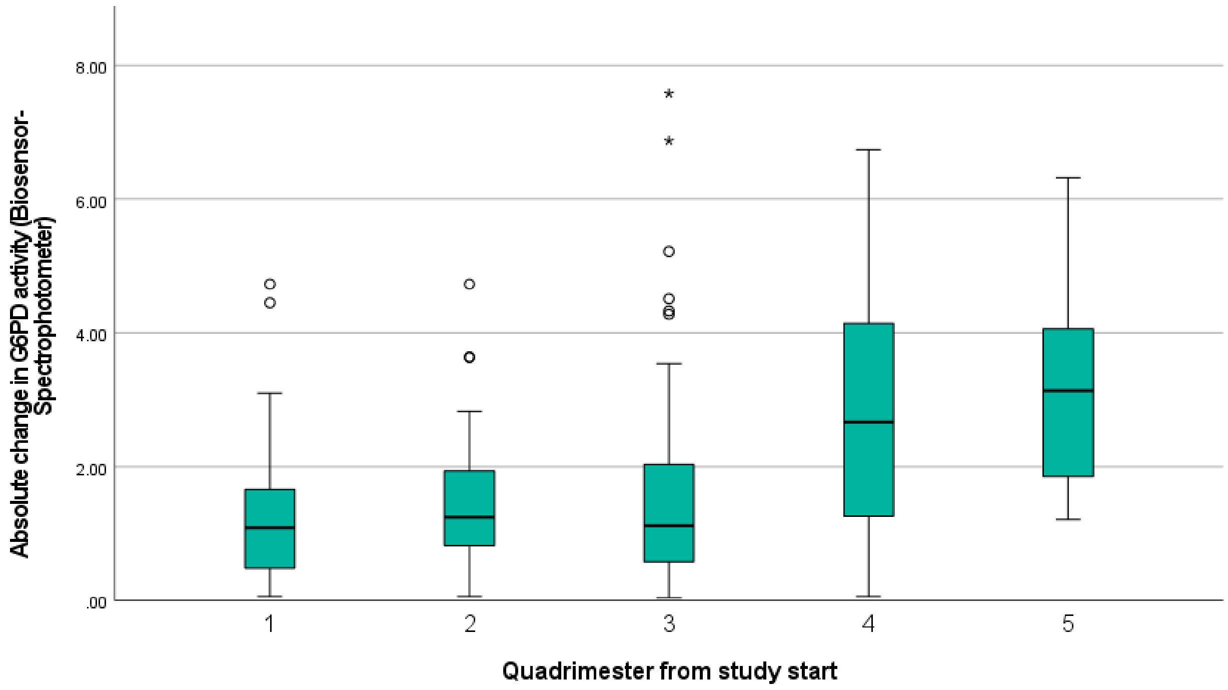

**S Table 3.** Characteristics of samples misclassified by Biosensor

| Clinic | Year | EGA | Sex | Reference G6PD (IU/gHb) | Reference Hb (g/dL) | Percent activity of reference (%) | Reference phenotype | Biosensor G6PD (IU/gHb) | Biosensor Hb (g/dL) | Percent activity of Biosensor (%) | Percent activity of reference (%) | Biosensor phenotype | G6PD genotype Mahidol | Retics (%) | WBC ($10^3$/ uL) |
|---|---|---|---|---|---|---|---|---|---|---|---|---|---|---|---|
| MKT | 2020 | 42 | F | 4.4 | 15 | 33 | INT | 4.3 | 15.2 | 30 | 32 | DEF | Heterozygote | 1.5 | 13.6 |
| MKT | 2021 | 40 | F | 7.1 | 15.8 | 54 | INT | 12.6 | 16.3 | 88 | 95 | NOR | Heterozygote | 1.3 | 20.2 |
| MKT | 2021 | 41 | F | 7.7 | 14.1 | 58 | INT | 12.5 | 11.5 | 87 | 94 | NOR | Heterozygote | ND | 20.6 |
| MKT | 2021 | 39 | F | 8.0 | 15.3 | 60 | INT | 10.9 | 11.7 | 76 | 82 | NOR | WT | 2.3 | 19.3 |
| MKT | 2021 | 39 | F | 8.2 | 14.3 | 62 | INT | 4 | 15.7 | 28 | 30 | DEF | Heterozygote | 2.2 | 21.1 |
| MKT | 2021 | 39 | F | 8.5 | 14.1 | 64 | INT | 11.8 | 14.1 | 82 | 89 | NOR | Heterozygote | ND | ND |
| WPA | 2021 | 39 | F | 9.4 | 13.3 | 71 | NOR | 9.8 | 13.7 | 68 | 74 | INT | Heterozygote | 1.6 | 13.8 |
| WPA | 2021 | 38 | F | 10.2 | 14.8 | 77 | NOR | 7.5 | 15.3 | 52 | 56 | INT | Heterozygote | 2.1 | 11.6 |
| WPA | 2021 | 39 | F | 10.9 | 15.6 | 82 | NOR | 8.8 | 16.7 | 61 | 66 | INT | WT | 1.6 | 12.5 |
| MKT | 2020 | 38 | F | 11.4 | 16.7 | 86 | NOR | 9.3 | 18.8 | 65 | 70 | INT | WT | 4.8 | 7.1 |
| WPA | 2021 | 39 | F | 11.7 | 14.3 | 88 | NOR | 8.1 | 16.5 | 56 | 61 | INT | WT | 1.8 | 11.1 |
| WPA | 2020 | 39 | F | 11.8 | 15.8 | 89 | NOR | 9.8 | 15.3 | 68 | 74 | INT | WT | 1.9 | 14.3 |
| MKT | 2020 | 39 | F | 12.1 | 12.6 | 91 | NOR | 9 | 13.2 | 63 | 68 | INT | WT | 3.9 | 14.4 |
| MKT | 2021 | 40 | F | 14.2 | 14 | 107 | NOR | 9.7 | 16.3 | 67 | 73 | INT | WT | 1.7 | 11.3 |
| MKT | 2021 | 37 | F | 15.1 | 11.2 | 113 | NOR | 8.2 | 12.1 | 57 | 62 | INT | WT | 3.7 | 15.5 |

**S Table 4.** Phototherapy treatment in newborns with EGA≥38 weeks with different G6PD phenotypes

| G6PD phenotype by spectrophotometry | PT | No PT | % PT | RR | 95%CI | $P_{Fisher}$ |
|---|---|---|---|---|---|---|
| Deficient | 8 | 20 | 28.6 | 4.9 | 2.3-10.5 | <0.001 |
| Intermediate | 3 | 17 | 15.0 | 2.6 | 0.8-8.1 | 0.13 |
| Normal | 15 | 242 | 5.8 | | | reference |
| G6PD phenotype by Biosensor | | | | | | |
| Deficient | 9 | 21 | 30.0 | 5.4 | 2.5-11.6 | <0.001 |
| Intermediate | 2 | 20 | 9.1 | 1.7 | 0.4-6.8 | 0.49 |
| Normal | 13 | 223 | 5.5 | | | reference |
