## Appendix 1 for "Quantitative G6PD point-of-care test can be used reliably on cord blood to identify male and female newborns at increased risk of neonatal hyperbilirubinaemia: a mixed method study"

SMRU
Shoklo Malaria Research Unit

### SD G6PD BIOSENSOR (for sample)

#### Prepare the machine, test device and buffer (step 1-8) BEFORE doing the blood collection (step 9)

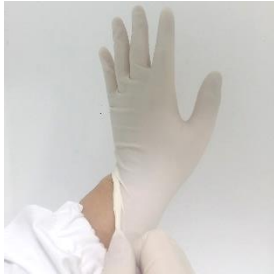

1. Put on gloves

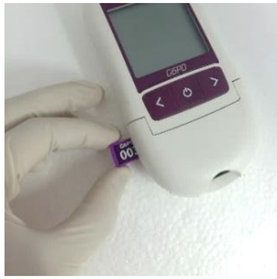

2. Insert codechip (For first time using or open new box of test device)

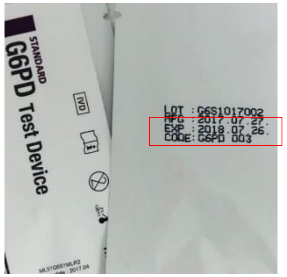


3. Check the expiry date printed on the foil pouch

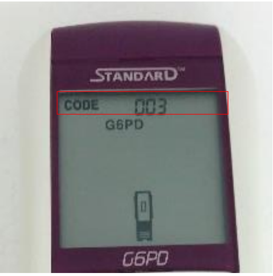


4. Check that codechip number on screen correspond to test device

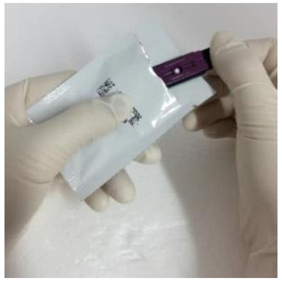

5. Open the foil pouch and take a test device out and hold the test in the right side

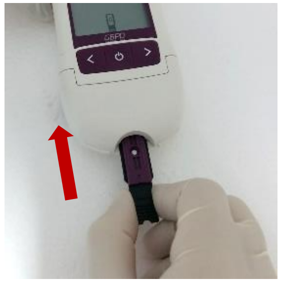

6. Insert the test device

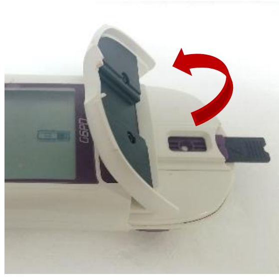

7. Open flap chamber

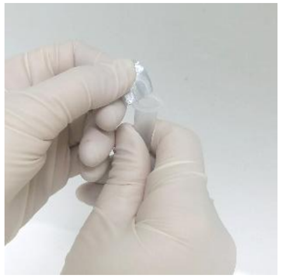

8. Open buffer tube and place on rack

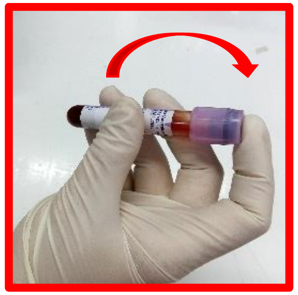

9. Mix sample tube well by inverting* 10 times
*Gently, no bubbles and no shaking

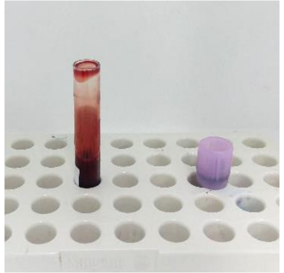

10. Place on rack

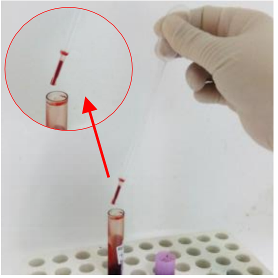

11. Collect blood by using Pasteur pipette

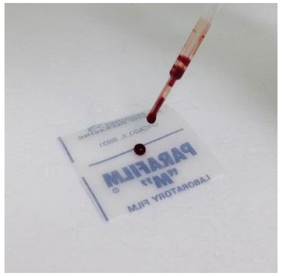

12. Drop blood on para film one drop (Avoid to make bubble)

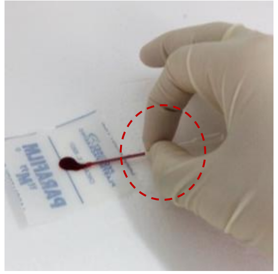

13. Hold the EZI tube horizontally, and touch the tip of the EZI tube to the blood specimen. Do not close hole.

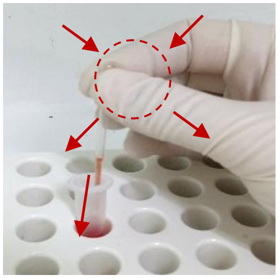

14. Mix the blood specimen with extraction buffer by pressing and releasing the EZI tube 10 times

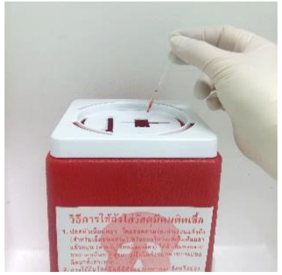

15. **Discard used EZI tube in the sharp bin**

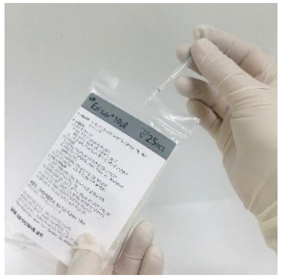

16. Take new EZI tube

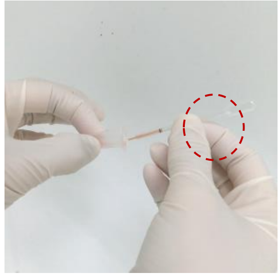

17. Hold the EZI tube horizontally, and touch the tip of the EZI tube to the mixed blood specimen. Do not close hole.

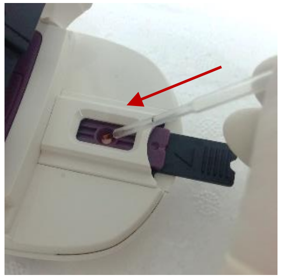

18. Apply mixed specimen to the specimen application hole of the test device

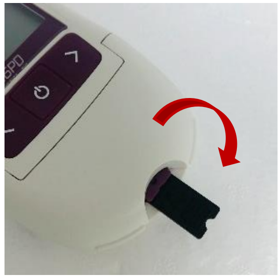

19. Close the flap chamber immediately after applying

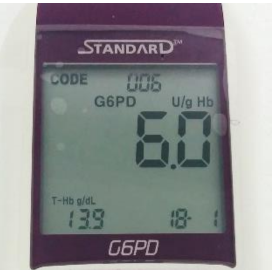


20. Wait for 2 min for the test result to appear on the screen (Check date) and report results on the logbook

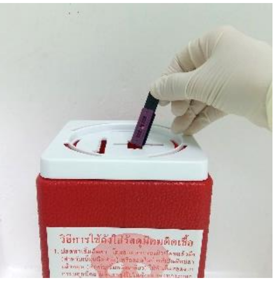

21. Take the used test device out and discard in sharp bin
